# Superiority of serum NFL in predicting Multiple Sclerosis severity

**DOI:** 10.1101/2021.05.07.21256836

**Authors:** Peter Kosa, Ruturaj Masvekar, Mika Komori, Jonathan Phillips, Vighnesh Ramesh, Mihael Varosanec, Mary Sandford, Bibiana Bielekova

**Affiliations:** Neuroimmunological Diseases Section, Laboratory of Clinical Immunology and Microbiology, National Institute of Allergy and Infectious Diseases, National Institutes of Health, Bethesda, MD, USA

**Author notes:** These authors contributed equally and are listed in alphabetical order (Peter Kosa, Ruturaj Masvekar). M.K. contributed to this work as a former employee of the National Institutes of Health, and the opinions expressed in this work do not represent her current affiliation (Eli Lilly Japan K.K., Kobe, Japan).

## Abstract

**Objective:** Serum neurofilament light chain (sNFL) is becoming an important biomarker of neuroaxonal injury. While sNFL correlates with cerebrospinal fluid NFL (cNFL), 40-60% of variance remains unexplained. Assuming that for diseases of the central nervous system (CNS), such as multiple sclerosis (MS), the cNFL better reflects CNS injury, our goal was to develop and validate adjustment of sNFL for relevant confounders, to strengthen cNFL-sNFL correlations.

**Methods:** We used 1,378 matched cNFL-sNFL pairs divided into training and validation cohort with matching data on 11 confounders, neuroexam, and magnetic resonance imaging (MRI). The effect of confounders on cNFL-sNFL relationship was tested using multiple linear regression (MLR) model. Propensity score matching was used to identify effect of spinal cord damage on sNFL levels.

**Results:** In the training cohort (n=898) we correlated 11 confounders with the residuals from cNFL-sNFL linear regression. Four non-overlapping confounders explaining highest proportion of variance (12%: age, 8.7%: blood urea nitrogen, 3%: alkaline phosphatase, and 3.9%: weight) were used in MLR model. The model strengthened the cNFL-sNFL correlation from R^2^ = 0.52 to 0.64 in the independent validation cohort and strengthened correlation of adjusted sNFL with number of contrast-enhancing lesions (from R^2^ 0.11 to 0.18). However, only sNFL, but not cNFL correlated with MS severity outcomes. Using propensity score matching, we demonstrated that subjects with proportionally higher sNFL to cNFL levels have significantly higher clinical and radiological evidence of spinal cord injury.

**Interpretation:** Superiority of sNFL likely resides in the release of NFL from axons of lower motor or dorsal ganglia neurons directly to blood.

## Introduction

Serum neurofilament light chain (sNFL) measured by advanced immunoassays capable of measuring single molecules or fg/ml concentrations (e.g., Single Molecule Array [SIMOA®]), is rapidly becoming an important minimally invasive biomarker reflecting neuro-axonal injury in neurological diseases, affecting both central (CNS) or peripheral (PNS) nervous systems.

NFL is one of the several structural axonal proteins, released during neuroaxonal injury. However, in contrast to other biomarkers of neuronal injury, such as ubiquitin carboxy-terminal hydrolase L1 (UCH-L1) and neuron-specific enolase (NSE), NFL has highly prolonged release after acute injury. For example, after traumatic brain injury (TBI), the serum levels of UCH-L1 and NSE peak at 24h and become undetectable 96 hours later ^1^. In contrast, NFL levels rise gradually, peak in several days and then remain elevated for up to a few months. While the gradual rise after TBI can be explained by Wallerian degeneration, the mechanism for prolonged increase after acute CNS injury, observed in many different diseases, is currently unexplained. Because the clearance of other CNS proteins (e.g., NSE and UCH-L1) from either cerebrospinal fluid (CSF) or blood are not affected by these neurological conditions, persistent NFL increase suggests its continuous release from axons for weeks/months after injury, perhaps due to remodeling of axonal/dendritic networks.

At any rate, this unusually prolonged increase of NFL after nervous system injury enhances its clinical utility, as it eliminates the test-timing problem: as long as the NFL is sampled within days/few weeks of injury, its levels correctly reflect original axonal damage.

Studies that directly compared sNFL with CSF NFL (cNFL) concentrations in the same subjects demonstrated that cNFL exerts stronger correlation with markers of acute CNS injury, such as brain MRI contrast-enhancing lesions (CEL), or by clinical correlates of acute CNS injury, such as multiple sclerosis (MS) relapses ^2^. Indeed, cNFL-sNFL correlations explain only about 60% of the measurement variance^3-5^. By focusing our work on CSF biomarkers, we were in a unique position to map biological contributors to the sources of this variance using matched CSF-serum samples. Consequently, the goal of this study was to develop (and validate in the independent cohort) a mathematical adjustment for relevant confounders that would better approximate sNFL to cNFL values. We expected that this will elucidate the mechanisms of sNFL clearance from the blood and enhance the clinical utility of sNFL test.

While we achieved our goal and demonstrated that sNFL adjusted for confounders better predicted presence of CEL, only sNFL, but not cNFL predicted MS severity, albeit weakly. Using propensity score matching, we showed that this advantage of sNFL resides, at least partially, in likely release of NFL directly to blood after the death of spinal cord neurons and associated peripheral nerve axons.

## Methods

### Subjects

1,378 matching CSF and serum samples were prospectively collected from 647 subjects from 7 diagnostic categories - Healthy Donors (HD), Relapsing-Remitting Multiple Sclerosis (RRMS), Primary Progressive MS (PPMS), Secondary Progressive MS (SPMS), Clinically Isolated Syndrome (CIS), Non-Inflammatory Neurological Diseases (NIND), and Other Inflammatory Neurological Diseases (OIND) as part of a Natural History protocol “Comprehensive Multimodal Analysis of Neuroimmunological Diseases of the Central Nervous System” (ClinicalTrials.gov Identifier: NCT00794352) or as part of the “NIB Repository Protocol” (10-N-0210). The protocols were approved by the NIH Institutional Review Board. All participants signed a written consent. Collected samples were split into training (2/3) and validation (1/3) cohort, controlling for diagnoses and keeping longitudinal samples in the same cohort to assure complete independence of the subjects in two cohorts. Demographic data are displayed in Table 1.

**Table 1:** Demographic data

|  | **Training cohort (n=424)** |  |  |  |  |  |  |
| --- | --- | --- | --- | --- | --- | --- | --- |
| Diagnosis | *CIS* | *HD* | *NIND* | *OIND* | *PPMS* | *RRMS* | *SPMS* |
| N (F/M) | 11/4 | 18/12 | 51/27 | 27/39 | 38/43 | 56/41 | 38/19 |
| Average age* (SD) | 41.1 (12.9) | 36.8 (13.4) | 48.5 (14.0) | 48.9 (14.4) | 54.0 (9.31) | 39.3 (9.82) | 51.5 (10.2) |
| Range (min-max) | 20.8-64.2 | 19.7-71.3 | 19.3-73.4 | 18.0-75.6 | 25.3-74.7 | 18.3-67.8 | 22.0-69.6 |
| N matching CSF - serum samples | 21 | 48 | 93 | 120 | 247 | 210 | 159 |
| Average # samples per subject (SD) | 1.4 (0.6) | 1.6 (1.0) | 1.2 (0.5) | 1.8 (1.6) | 3.1 (1.9) | 2.2 (1.6) | 2.8 (2.0) |
| Range (min-max) | 1-3 | 1-4 | 1-3 | 1-9 | 1-8 | 1-9 | 1-7 |
|  | **Validation cohort (n=223)** |  |  |  |  |  |  |
| Diagnosis | *CIS* | *HD* | *NIND* | *OIND* | *PPMS* | *RRMS* | *SPMS* |
| N (F/M) | 6/3 | 6/11 | 29/12 | 13/21 | 23/19 | 33/17 | 17/13 |
| Average age* (SD) | 41.7 (11.1) | 39.1 (13.7) | 47.9 (11.6) | 42.4 (15.5) | 54.4 (9.0) | 43.4 (11.4) | 51.1 (10.6) |
| Range (min-max) | 26.7-57.7 | 19.4-56.8 | 18.2-74.9 | 15.8-68.8 | 28.2-65.8 | 18.0-68.6 | 27.4-68.0 |
| N matching CSF - serum samples | 13 | 26 | 48 | 65 | 142 | 92 | 94 |
| Average N samples per subject (SD) | 1.4 (0.7) | 1.5 (0.9) | 1.2 (0.6) | 1.9 (1.8) | 3.4 (1.8) | 1.8 (1.3) | 3.1 (1.9) |
| Range (min-max) | 1-3 | 1-4 | 1-4 | 1-8 | 1-7 | 1-6 | 1-8 |

\* age at the first collection date per subject
F – female, M – male, N – number, min – minimum, max – maximum, SD – standard deviation

A set of body measures was taken, and laboratory tests were performed at the time of CSF/blood collection at the NIH Department of Laboratory Medicine and recorded in the NIH electronical medical record system. MS patients underwent a full neurological exam at the time of sample collection that was documented electronically using NeurEx™ App ^6^ and a research brain MRI (with or without gadolinium contrast) that was reviewed and graded by a board-certified neurologist and recorded using COMRIS tool ^7^ in our research database. MS severity outcomes were calculated as described ^8-10^. All laboratory, clinical, and MRI outcomes were collected and generated prior to evaluation of NFL levels in CSF and serum samples.

### Sample collection

Samples were collected following the laboratory Standard Operating Procedures. Briefly, CSF, collected by lumbar puncture, was kept on ice upon collection and processed withing 15 min of collection by centrifugation at 335g for 10 min at 4°C; the supernatant was aliquoted and stored at -80°C. Blood was collected by venipuncture using the SST tube, incubated at room temperature for 30 min, then spun at 2,000g for 10 min at 4°C. The serum was aliquoted and stored at -80°C. Personnel processing samples were blinded to patients’ diagnoses/clinical/MRI outcomes.

### NFL ELISA

NFL concentration in CSF samples were measured using a solid-phase sandwich ELISA kit (UmanDiagnostics, Umea, Sweden; Catalog number: 10-7002 RUO; Lower Limit of Detection (LLoD): 33 pg/ml); assay was performed as per manufacturer’s instructions. Briefly, all samples were diluted 1:2 with provided sample diluent and then analyzed blindly and in singlets. Samples were analyzed on multiple plates; location of samples on each plate was randomized and a control sample was analyzed in duplicate. The coefficient of variance (CV) for the control sample across the plates was <20%, confirming the assay precision and reproducibility.

### NFL Single Molecule Array (Simoa™) Assay

NFL levels in serum samples were analyzed using a Simoa™ assay kit (Quanterix, Billerica, MA, USA; Product number: 103186; LLoD: 0.038 pg/ml) on Simoa HD-1 analyzer; assay was performed as per manufacturer’s instructions. Briefly, all samples were diluted 1:4 with provided sample diluent using on board dilution functionality, and then analyzed blindly in singlets. Samples were analyzed in multiple batches; each plate contained the quality control (QC) samples provided with kit. The QC samples had measured concentrations within acceptable range, confirming the assay precision.

### Statistics

All modeling and analyses were performed in the training cohort, the resulting models/equations were then tested in the independent validation cohort. All analyses and plots were generated in R Studio Version 1.1.463 (R version 4.0.2) ^11^. Correlations between variables were assessed using *lm* function (“stats” package), generating Coefficient of variance (R^2^) and p-value. Final multiple linear regression model was selected using stepwise algorithm in *stepAIC* function (“MASS” package^12^). Propensity score matching was performed using *matchit* function with “full” method (“MatchIt” package^13^). Differences between propensity score-matched groups were evaluated by *stat_compare_means* function (“ggpubr” package ^14^) using paired *wilcox*.*test* or *t*.*test* method.

## Results

### Assessing univariate correlations of 11 candidate confounding factors with cNFL-sNFL residuals in the training cohort

We measured NFL levels in a cohort of 1,387 matching CSF and serum samples consisting of 7 diagnostic groups (HD, NIND, OIND, CIS, RRMS, PPMS, and SPMS). This cohort was split into training (2/3) and validation (1/3) dataset prior to running any analysis, balancing for the diagnosis. The correlation between sNFL and cNFL in the training cohort showed that sNFL explains 53% of variance of cNFL (Fig 1A). Calculation of residuals from this linear regression model (Fig 1B,C) removed the 53% of variance explained by sNFL and allowed us to identify additional confounding factors that would explain additional variance observed between sNFL and cNFL. Correlation analysis of NFL residuals with 11 variables reflecting aging (age), distribution volume (Body Mass Index [BMI], height, weight, estimated blood volume [eBV]), liver and kidney functions (serum aspartame aminotransferase [AST], serum alanine aminotransferase [ALT], Blood Urea Nitrogen [BUN], serum creatinine, and estimated Glomerular Filtration Rate [eGFR]), and reticuloendothelial system (serum alkaline phosphatase [AP]) (Fig 1D) identified four non-overlapping variables with statistically significant correlation with NFL residuals, explaining 12% of variance for age, 8.7% for BUN, 3.9% for weight, and 3% for AP (Fig 1E). Using stepwise regression, we tested whether addition of the four identified variables improves the performance of a multiple linear regression (MLR) model predicting sNFL levels from cNFL levels. In the training cohort, all four variables contributed significantly to improvement of the MLR model performance, with weight having the strongest influence (t value -8.03), followed by age (t value 7.74), BUN (t value 6.79), and AP (t value 3.22).

**Figure 1:**
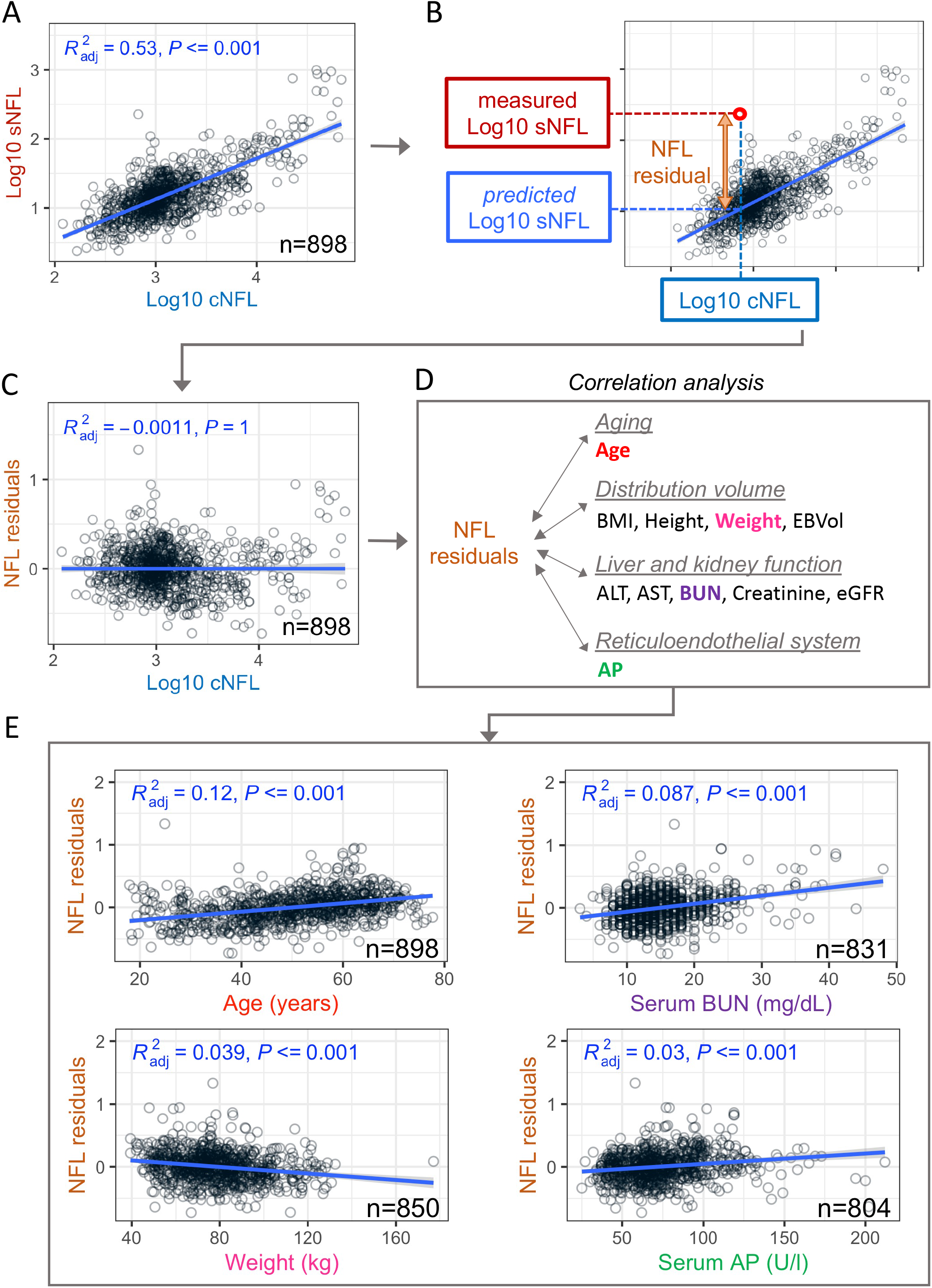
Variance between sNFL and cNFL concentrations. (A) Linear regression model between log10-transformed concentration (pg/ml) of sNFL and cNFL in the training cohort of samples. (B) Generation of NFL residuals (orange) as differences between measured sNFL concentration (in red) and predicted sNFL concentration (in light blue) calculated from measured cNFL (dark blue) using linear regression model. (C) Correlation between NFL residuals (y-axis) and log10 cNFL concentration (x-axis) explains 0% of variance. (D) Generated NFL residuals were correlated with 11 potential confounders representing aging, distribution volume, liver and kidney function, and reticuloendothelial system. (E) Four non-overlapping outcomes show statistically significant correlation with NFL residuals, explaining 3-12% of variance. Blue line represents linear regression model with gray shading corresponding to 95% confidence interval. BMI – body mass index, EBVol – estimated blood volume (calculated using Nadler’s equation ^19^), ALT – alanine aminotransferase, AST – aspartame aminotransferase, BUN – blood urea nitrogen, eGFR – estimated glomerular filtration rate (calculated using 4-variable MDRD Study equation ^20^), AP – alkaline phosphatase. The attrition of sample number was caused by exclusion of samples with missing data.

### Adjusting sNFL levels for four confounding factors using multiple linear regression model developed in the training cohort, strengthens correlations between sNFL and cNFL in the independent validation cohort

Addition of age, weight, BUN, and AP as explanatory variables into a MLR model improved the predictive power of cNFL in the training cohort from 56% of sNFL variance explained in the simple linear regression model (Fig 2A) to 66% of variance explained in the MLR model (Fig 2B). The performance of this MLR model was evaluated in an independent validation cohort, where the addition of the four explanatory variables increased the % of variance explained from 52% in the simple linear regression model (Fig 2C) to 64% in the MLR model (Fig 2D). The final MLR equation is as follows:

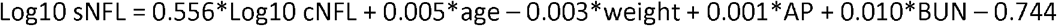

**Figure 2:**
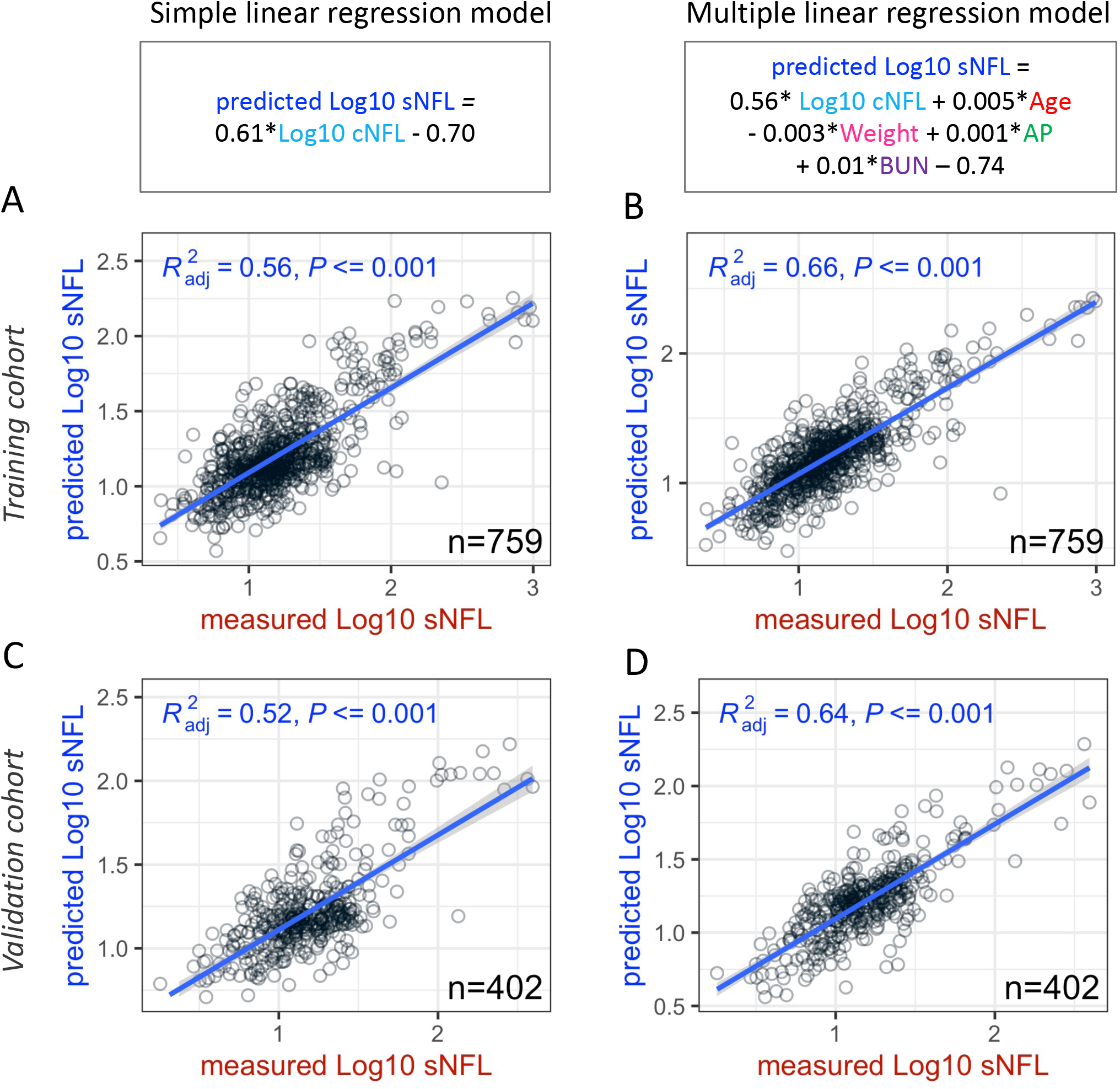
Multiple linear regression model improves correlation between cNFL and sNFL. Correlation analysis of measured sNFL (x-axes) and predicted sNFL (y-axes) in training (A,B) and validation (C,D) cohorts shows improved correlation when multiple linear regression model, including age, weight, alkaline phosphatase (AP) and blood urea nitrogen (BUN), is used for predicting sNFL (B,D) instead of simple linear regression model (A,C). Blue line represents linear regression model with gray shading corresponding to 95% confidence interval. The attrition of sample number in both training and validation cohort was caused by exclusion of samples with missing data.

### Adjusted sNFL correlates better with MRI Contrast-Enhancing Lesions (CEL) than un-adjusted sNFL (but weaker than cNFL)

Although sNFL and cNFL correlate strongly, the CNS tissue destruction reflected by the number of MRI CEL can be predicted more accurately using cNFL, rather than sNFL; cNFL explains 24% of variance of CEL, while sNFL only 14% in the training cohort (Fig 3A,B) and similar correlation can be observed in the validation (20% of variance explained by cNFL versus 11% of variance explained by sNFL) (Fig 3D,E). We asked whether adjustment of sNFL for four identified confounders would improve its correlation with the CEL. Therefore, we reshuffled the MLR equation to predict cNFL from sNFL levels as follows:

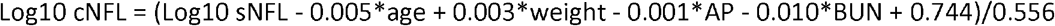

**Figure 3:**
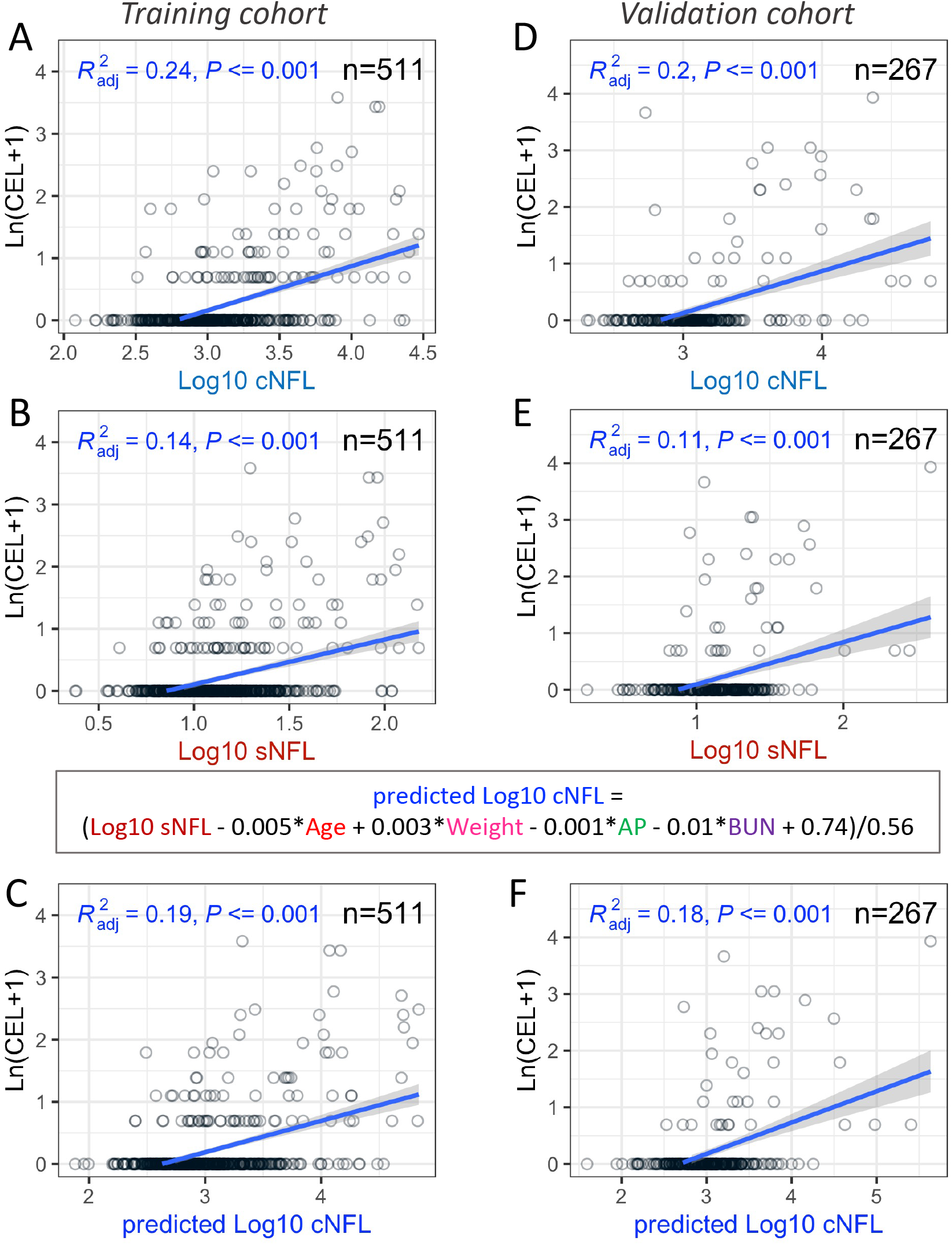
Adjustment for four confounder improves correlation of sNFL with number of MRI contrast enhancing lesions (CEL) and eliminates noise. Correlation analysis between CEL (y-axes - transformed as natural logarithm of [CEL+1]) and NFL (x-axes) shows higher predictive power of cNFL in both training (A) and validation (B) cohort, compared to sNFL in training (C) and validation (D) cohort. Adjustment of sNFL for four confounders (age, weight, alkaline phosphatase [AP] and blood urea nitrogen [BUN]) increases the correlation with CEL in both training (E) and validation (F) cohort compared to unadjusted sNFL. Blue line represents linear regression model with gray shading corresponding to 95% confidence interval. The attrition of sample number in both training and validation cohort was caused by exclusion of samples with missing data.

The predicted cNFL showed increased correlation with CEL in comparison to sNFL, explaining 19% of variance in the training cohort and 18% of variance in the validation cohort (Fig 3C,F).

This validated the hypothesis that cNFL is better marker of acute CNS injury (measured in MS by CEL) and that adjusting sNFL levels for covariates that affect distribution and metabolism of cNFL released to blood, strengthens the sNFL ability to predict acute CNS injury.

### sNFL, but not cNFL correlates with MS severity outcomes

While CEL is a biomarker of acute CNS injury in MS, it correlates poorly with the rates of disability progression. Thus, to ask whether cNFL, sNFL and adjusted sNFL have clinical value in patients who may not form new MS lesions, we correlated these biomarkers with MS disease severity outcomes. Correlation of Multiple Sclerosis Severity Scale (MSSS) ^9^, Age-Related Multiple Sclerosis Severity (ARMSS)^8^, and Multiple Sclerosis Disease Severity Scale (MS-DSS) ^10^ showed that sNFL consistently outperforms cNFL (Fig 4). None of the severity outcomes correlated significantly with cNFL levels, but all three of them showed statistically significant correlation with sNFL, explaining 1.6% (MS-DSS), 2.8% (ARMSS), and 5.7% (MSSS) of variance (Fig 4A). Similar results were observed in the validation cohort, although the variance explained by sNFL was higher (8.9% for MS-DSS, 5.9% for ARMSS, and 14% for MSSS) (Fig 4B). Thus, we conclude that sNFL, but not cNFL reproducibly correlates with MS severity outcomes, although it explains less than 15% of variance.

**Figure 4:**
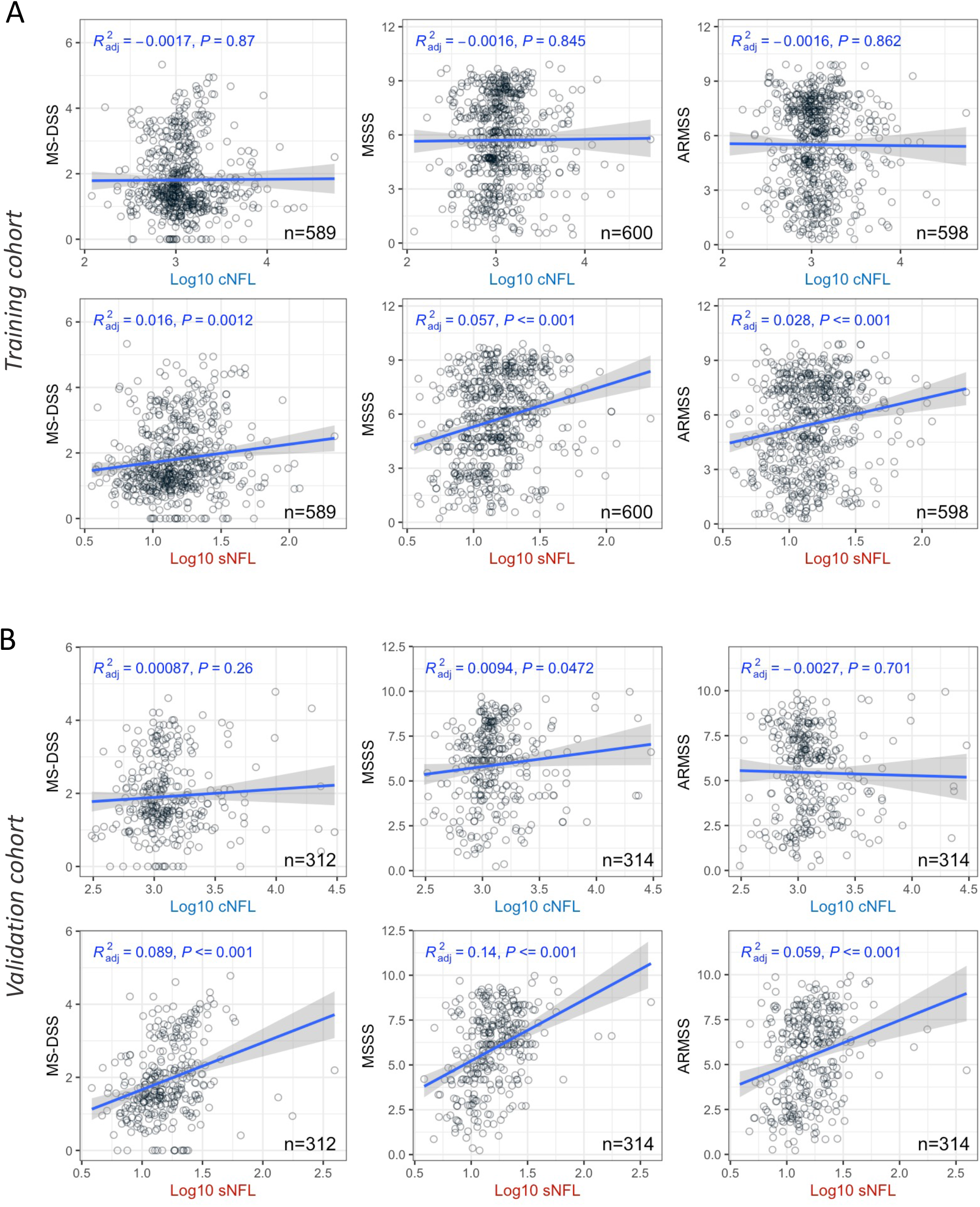
sNFL correlates better with MS disease severity outcomes than cNFL. Correlation analysis of three MS severity outcomes - Multiple Sclerosis Disease Severity Scale (MS-DSS), Multiple Sclerosis Severity Score (MSSS), and Age Related Multiple Sclerosis Severity Score (ARMSS) – with sNFL and cNFL levels shows higher proportion of variance explained by sNFL level than cNFL levels in both training (A) and validation (B) cohort of MS subjects only. Blue line represents linear regression model with gray shading corresponding to 95% confidence interval. The attrition of sample number in both training and validation cohort was caused by exclusion of samples with missing data.

Because MSSS consistently outperformed ARMSS and MS-DSS (both of which are adjusted for the age of the patient in contrast to MSSS) in the univariate correlations of measured cNFL and sNFL with MS outcomes, age likely underlies at least some of this sNFL advantage. Indeed, while both cNFL and sNFL increase with age, age also explains 12% of variance of sNFL-cNFL residuals (Fig. 1E), which means that sNFL increases with age more robustly than cNFL. However, which aspects of aging, or of MS evolution that occurs with aging, explains this advantage of sNFL over cNFL?

**Testing two mutually non-exclusive hypotheses in the training cohort that may explain superiority of sNFL for MS severity outcomes: 1. Diluting of cNFL in patients with brain atrophy and 2. Increasing sNFL by direct release of NFL to blood in spinal cord injury**

We generated and tested two mutually non-exclusive hypotheses that could explain why sNFL better predicts MS severity: 1) increased brain atrophy that is associated with aging of MS patients leads to dilution of cNFL, resulting in proportionally higher concentration of sNFL, 2) increase in spinal cord (SC) injury that has been associated with aging and more severe disease leads to direct release of NFL into blood.

We tested these hypotheses using propensity score matching (Fig 5) to maximize differences in appropriate cNFL-sNFL residuals: For the effect of brain atrophy, we selected subjects matched for sNFL levels but with vastly different cNFL levels (Fig 6), while for testing the possible release of NFL from spinal cord injury directly to serum we selected subjects matched for their cNFL levels, but with very different sNFL (Fig 7).

**Figure 5:**
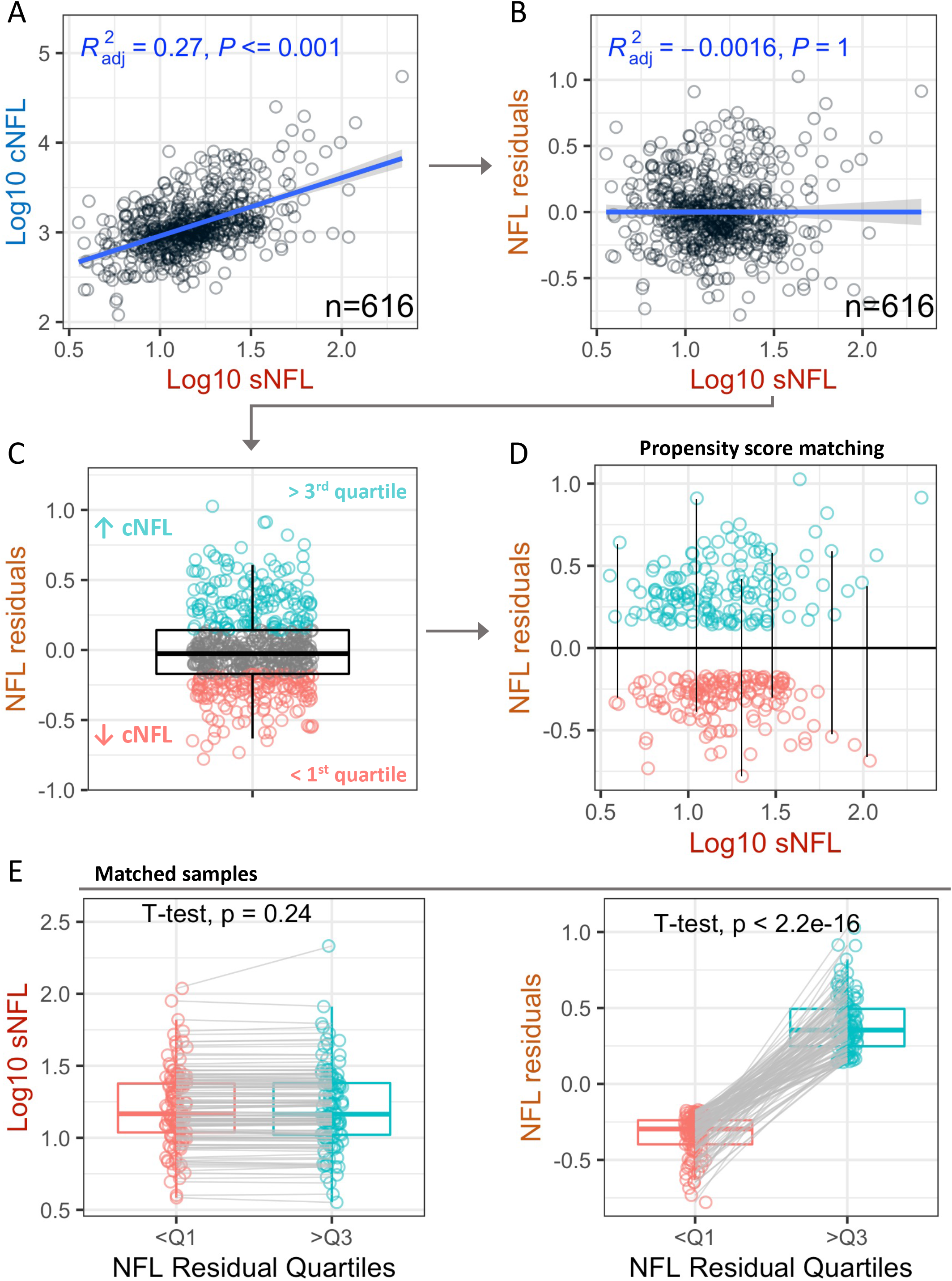
Propensity score matching of samples with proportionally different levels of NFL residuals. (A) In the training cohort of MS samples a linear regression model of cNFL versus sNFL was generated and (B) used to calculate NFL residuals. (C) The 1^st^ and 3^rd^ quartile of NFL residual values was determined and used to generate two groups of samples: sample with NFL residuals below the 1^st^ quartile (salmon – samples with measured cNFL levels lower than predicted by linear regression model) and samples with NFL residuals above the 3^rd^ quartile (light cyan - samples with measured cNFL levels higher than predicted by linear regression model). (D) Propensity score matching algorithm selects pairs of samples (black vertical lines) with close to identical levels of sNFL (x-axis), but significantly different levels of NFL residuals (y-axis). (E) In the matched dataset the paired T-test showed no statistically significant difference in sNFL levels between the two groups of samples (left), while there was statistically significant difference in NFL residual levels in the same set of paired samples (right).

**Figure 6:**
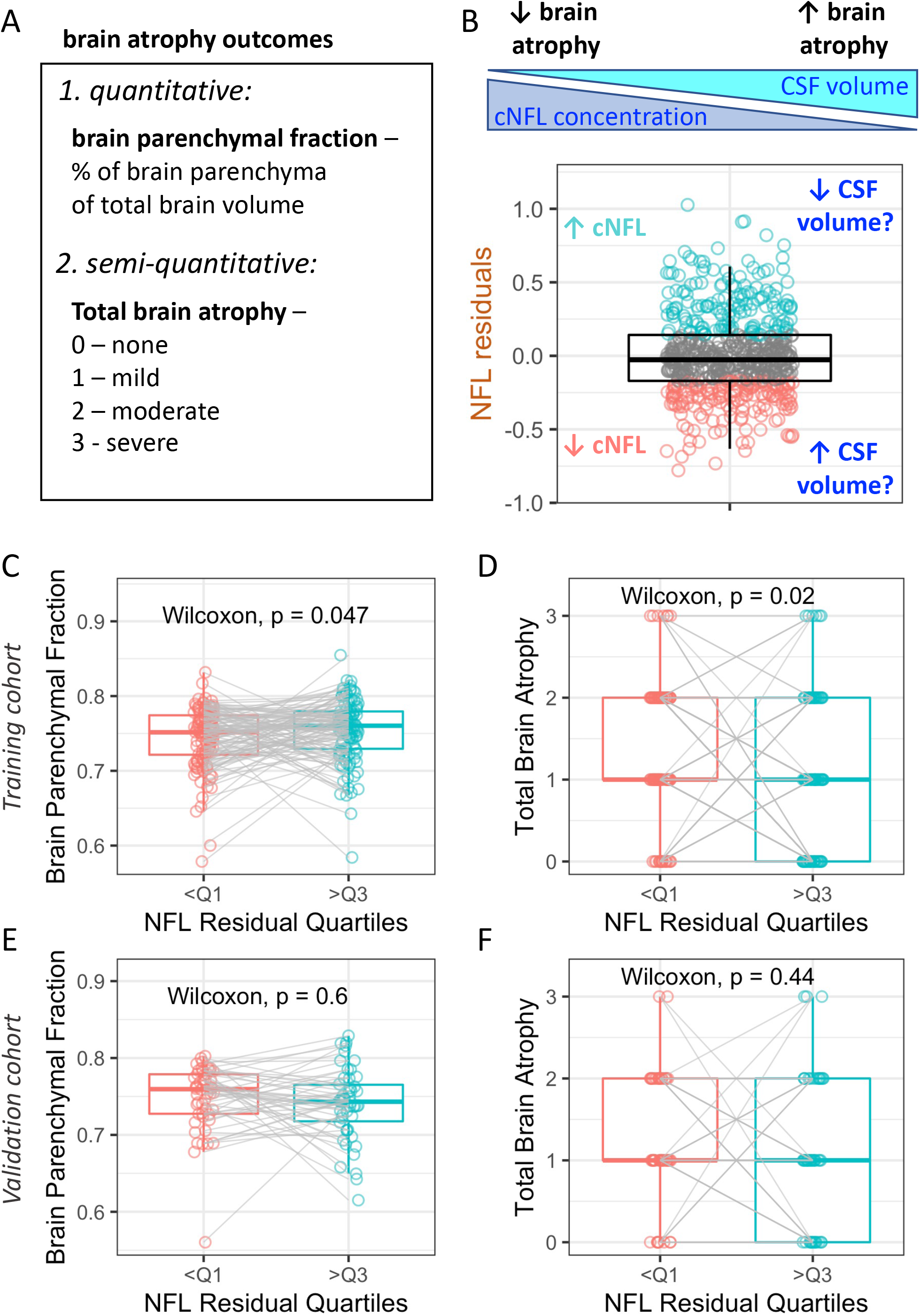
Brain atrophy does not explain the proportional differences in cNFL levels compared to sNFL levels. (A) Brain atrophy was evaluated by brain parenchymal fraction (a fully quantitative MRI outcome generated as a ratio of the sum of supratentorial gray matter, white matter, and lesions, divided but the volume of the brain tissue and cerebrospinal fluid) and by semi-quantitative outcome of total brain atrophy (visual grading of levels of global brain atrophy as none, mild, moderate, and severe). (B) The tested hypothesis assumes that higher brain atrophy that results in larger CSF volume would lead to dilution of cNFL leading to proportionally lower concentration in cNFL in subjects with higher brain atrophy. Paired Wilcoxon Ranked Sum Test showed statistically significant difference in both brain parenchymal fraction (C) and total brain atrophy (D) between samples with proportionally higher and lower cNFL levels in the training cohort. The observed differences were not confirmed in the independent validation cohort neither for brain parenchymal fraction (E) or total brain atrophy (F).

**Figure 7:**
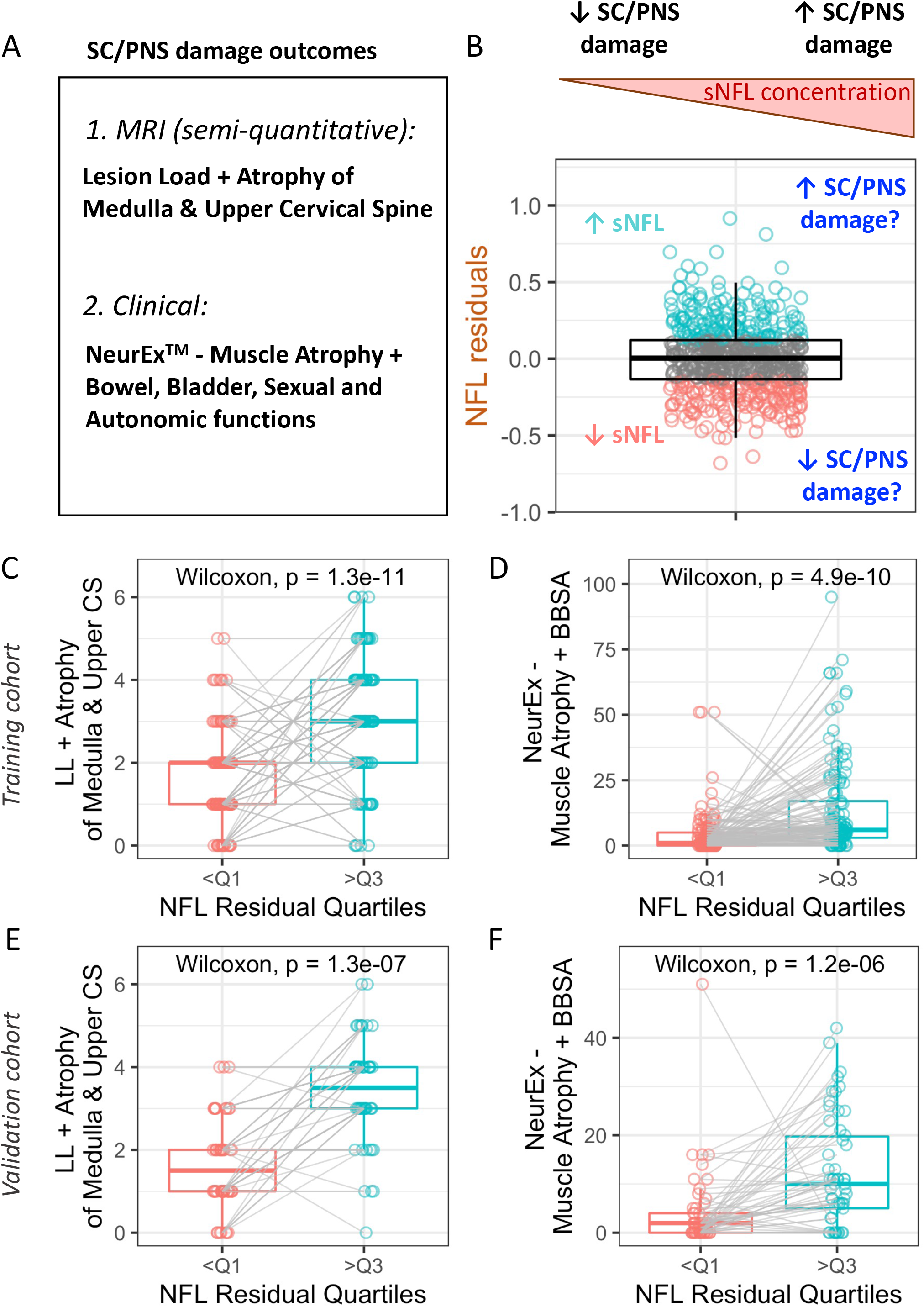
Spinal cord (SC) and/or peripheral nervous system (PNS) damage are associated with increased levels of sNFL. (A) SC/PNS damage was evaluated using a semi-quantitative MRI outcome – a sum of scores for lesion load (LL) and atrophy evaluated at the level of medulla and cervical spine (CS) and by clinical outcome generated from digitally documented neurological exam using NeurEx™ App as a sum of score for muscle atrophy (capturing damage of motor neurons) and score for bowel, bladder, sexual and autonomic dysfunctions (BBSA - capturing damage to peripheral/autonomous nervous system). (B) The tested hypothesis assumes that damage to SC/PNS results in direct release of NFL into blood and therefore subjects with higher SC/PNS damage will have proportionally elevated sNFL levels compared to cNFL levels. (C) Paired Wilcoxon Ranked Sum Test showed statistically significant difference in both MRI (C) and clinical (D) outcomes between samples with proportionally higher and lower sNFL levels in the training cohort. The observed differences were confirmed in the independent validation cohort for both MRI (E) and clinical (F) outcomes measuring the levels of SC/PNS damage.

We will illustrate the propensity score matching only for the first comparison, testing effect of brain atrophy: In the MS training cohort we calculated NFL residuals by removing the variance explained by sNFL (Fig 5A,B). Next, we calculated the first and the third quartile of the dataset distribution and isolated samples with NFL residuals below the 1^st^ quartile (samples with measured cNFL levels higher than predicted by the linear regression model) and samples above the 3^rd^ quartile (samples with measured cNFL levels higher than predicted by the linear regression model) (Fig 5C). Using propensity score matching (Fig 5D), we generate pairs of samples where the levels of sNFL were close to identical (Fig 5E left), while the levels of NFL residuals were statistically different (Fig 5E right). Such dataset allowed us to use more powerful statistical test on paired samples.

We tested two outcomes of brain atrophy (Fig 6A): 1) a fully quantitative measure called brain parenchymal fraction (BPFr), that compares the volume of brain gray matter, white matter, and lesions, to total brain volume (sum of brain tissue volume and CSF volume) and 2) semiquantitative measure generated by visual grading of the atrophy based on T1 and T2 brain MRI sequences into four categories (none, mild, moderate, and severe) ^7^. The amount of SC damage can be also quantified in two ways (Fig 7A)– based on MRI imaging, where semi-quantitative grading of lesion load and atrophy of medulla and upper CS is performed on brain MRI images, following the similar grading pattern of none, mild, moderate, and severe ^7^. The drawback of this measurement is that it does not capture the damage to the thoracic or lumbosacral spinal cord. Therefore, we sought to employ complementary information from the Neurological examination. NeurEx™ App – a digitalized neuroexam –allows granular measurements of neurological disability in different functional systems ^6^. Two parts of NeurEx™ can be used as surrogates of SC and peripheral nervous system (PNS) injury – muscle atrophy (which in MS patients is a consequence of damaged lower motor neurons in the spinal cord, axons of which constitute motor fibers of peripheral nerves), and bowel, bladder, sexual, and autonomic (BBSA) dysfunctions (in MS likely caused by death/injury of the autonomic neurons of the spinal cord that project axons to the peripherally-located autonomic ganglia).

Using these four outcomes we were able to test our two hypotheses by asking following questions: do patients with proportionally lower concentrations of cNFL compared to sNFL have increased brain atrophy (larger CSF volume) (Fig 6B), and analogously, do patients with proportionally higher concentrations of sNFL compared to cNFL have increased SC/PNS damage (Fig 7B)?

In the training cohort, we observed statistically significant differences in brain atrophy measured by both quantitative and semi-quantitative outcome between groups of samples with proportionally lower and higher cNFL levels compared to sNFL levels (Fig 6C,D). The observed differences were in agreement with the proposed model – samples with proportionally higher cNFL had significantly lower amount of brain atrophy measured by both MRI outcomes, but the effect sizes were small and the p-values were marginal.

We also observed statistically significant difference in both SC/PNS damage outcomes between groups of samples with proportionally higher and lower levels of sNFL compared to cNFL (Fig 7C,D). In both cases, the samples with proportionally higher sNFL were associated with higher levels of SC/PNS damage with very low p-values, fitting the proposed model.

### Validating spinal cord injury as the dominant reason for superiority of sNFL in predicting MS severity outcomes

We then generated analogous two propensity score-matched datasets in the new set of patients/samples (i.e., independent validation cohort) – one with groups of proportionally different cNFL levels and one with proportionally different sNFL levels to validate the training cohort observations.

Consistent with marginal p-values observed in larger training cohort, we were unable to validate the difference in either brain parenchymal fraction or semi-quantitative measure of brain atrophy in the (smaller) independent validation cohort (Fig 6E,F).

In contrast, both outcomes assessing SC/PNS damage by a semiquantitative measure of lesion load and atrophy of medulla and upper CS, as well as NeurEx™ outcome comprised of the level of muscle atrophy and BBSA dysfunctions robustly validated their statistically significant differences between samples with proportionally higher and lower sNFL levels compared to cNFL levels (Fig 7E,F).

Therefore, we conclude that SC damage underlies stronger predictive power of sNFL versus cNFL for MS severity outcomes. The most logical explanation of our validated observations is that SC injury to the lower motor neurons and the neurons of the sympathetic and parasympathetic nervous system results in direct or indirect (i.e., Wallerian) injury to their axons, which constitute peripheral nerves and which therefore release their NFL directly to blood, bypassing the CSF.

## Discussion

We started this work with the premise that cNFL is clinically more relevant biomarker for MS than sNFL and that we might enhance the clinical value of sNFL by adjusting for relevant confounders.

We validated this idea only partially: we identified reproducible confounders and validated a MLR model that adjusted sNFL to better approximate cNFL concentrations. Adjusted sNFL then demonstrated increased correlations with MRI CELs in MS patients.

Analyzing contributions to our multiple linear regression model, we conclude that kidney function represents the most important clearance mechanism for sNFL, followed by reticuloendothelial system. Dilution of sNFL based on the peripheral distribution volume reflected by weight played a minor role. The most surprising observation was that the strongest confounder was age, explaining 12% variance of the cNFL-sNFL residuals.

We knew that cNFL and sNFL concentrations increased with healthy aging ^4, 15^. However, the data presented here showed that this increase was more robust for sNFL than for cNFL. We considered the possibility of age-related dilution of cNFL in the increasing CSF volume. Assuming that both CSF production and its drainage to blood remain constant during aging, increased CSF volume (i.e., brain atrophy) would lead to lower steady state cNFL concentration. On the other hand, as systemic distribution volume remains constant with aging (on a group level), the sNFL concentrations would not be affected, leading to proportionally higher sNFL compared to cNFL concentrations. However, when we tested this hypothesis using propensity score matching, we did not see any effect of brain atrophy, whether using semi-quantitative or fully quantitative (i.e., BPFr) MRI biomarkers on cNFL-sNFL residuals.

We then considered the possibility that proportionally increased sNFL levels may be due to release of NFL from peripheral axons/nerves, directly to the blood. Indeed, dying SC neurons, which axons comprise peripheral nerves (i.e., lower motor neurons, sympathetic and parasympathetic neurons, dorsal root ganglion cells) would release, during degeneration of their axons, NFL to blood bypassing CSF. This led to intriguing hypothesis, that the higher sNFL residuals might reflect higher SC injury. Because SC injury poses worse prognosis in MS, leading to faster accumulation of physical disability^16^, this could fully explain the superiority of sNFL correlations with MS severity measures.

We already knew that subjects with peripheral nerve disorders (PNS), such as Guillain-Barre Syndrome (GBS), have proportionally higher sNFL to cNFL levels compared to subjects with CNS disorders such as Alzheimer’s disease, or compared to patients with CNS/PNS disease such as Amyotrophic lateral sclerosis (ALS) ^17^. While we did not collect any data that would indicate peripheral nerve injury, we measured SC injury using prospectively acquired imaging and clinical biomarkers. Performing extensive analyses in the training cohort on propensity score-matched samples that had identical cNFL concentrations, but significantly different sNFL levels we found, reassuringly, that all measures of SC injury were robustly elevated in the patients with proportionally higher sNFL concentrations. Consistent with very low p-values in the training cohort, the strong, positive associations between imaging and clinical measures of SC injury and higher sNFL residuals were easily validated in the new set of MS patients. We conclude that congruency of these measurements between the two independent cohorts provides strong support for the hypothesis that superiority of sNFL over cNFL in predicting MS severity resides in its ability to reflect, in addition to cNFL levels, also SC injury that increases only sNFL, but not cNFL concentrations.

The observations that cancer chemotherapy and environmental toxins cause nerve damage much more readily than CNS damage was interpreted as evidence that blood-nerve barrier, lacking astroglial processes and lamina limitans is more permeable to toxins than blood-brain barrier ^18^. If this is true, then aging processes should also lead to proportionally higher increase of NFL from PNS, rather than CNS tissue, which is exactly what we see in healthy subjects (age explains 15% of NFL residual variance in the HD cohort, p=0.004).

In conclusion, this study validated adjustments of sNFL for confounding factors, to increase precision in identifying MS patients with breakthrough lesional activity measured by CEL using minimally invasive blood biomarker. It also showed that sNFL (but not cNFL) values correlate with multiple MS severity measures, due to release of NFL from axons constituting peripheral nerves to blood, bypassing the CSF. This explains, at least in part, why sNFL increases more robustly with age than cNFL. We can generalize the new knowledge presented in this paper, that future ultrasensitive assays that measure blood analytes originating exclusively from neurons, and possibly also from myelin, may have better clinical value than their CSF counterparts in predicting MS severity, because blood measurement aggregate both sources of the analyte: CNS and PNS. In fact, measuring these analytes in both compartments provides an opportunity to identify (subclinical) SC or PNS injury and therefore may aid in the diagnostic process.

## Data Availability

All data referred to in the manuscript are available upon request.

## Acknowledgments

We would like to thank clinicians Alison Wichman and Jamie Cherup, research nurses Tiffany Hauser, Naomi Gathua, Jenifer Dwyer, and patient care coordinators Michelle Woodland, Kewonie Pumphrey, for their excellent patient care. We would like to thank laboratory technician Elena Romm for processing patient samples, Finally, we would like to thank our patients, their caregivers, and healthy volunteers for being partners in research – without them this work would be possible.

This study was funded by the Division of Intramural Research, National Institute of Allergy and Infectious Diseases, National Institutes of Health.

## Authors’ contributions

B.B. designed and supervised the study. P.K., R.M., M.K., J.P., V.R., M.V., M.S., B.B. contributed to acquisition and analysis of the data. B.B., P.K., R.M. drafted the text and prepared the figures.

